# Show Your Work: Verbatim Evidence Requirements and Automated Assessment for Large Language Models in Biomedical Text Processing

**DOI:** 10.64898/2026.03.03.26346690

**Authors:** Paul Windisch, Julia Weyrich, Fabio Dennstädt, Daniel R. Zwahlen, Robert Förster, Christina Schröder

## Abstract

**Purpose:** Large language models (LLMs) are used for biomedical text processing, but individual decisions are often hard to audit. We evaluated whether enforcing a mechanically checkable “show your work” quote affects accuracy, stability, and verifiability for trial eligibility-scope classification from abstracts.

**Methods:** We used 200 oncology randomized controlled trials (2005 - 2023) and provided models with only the title and abstract. Trials were labeled with whether they allowed for the inclusion of patients with localized and/or metastatic disease. Three flagship models (GPT-5.2, Gemini 3 Flash, Claude Opus 4.5) were queried with default settings in two independent conditions: label-only and label plus a verbatim supporting quote. Models could abstain if they deemed the abstract to not contain sufficient information. Each condition was repeated three times per abstract. Quotes were mechanically validated as exact substrings after whitespace normalization, and a separate judge step used an LLM to rate whether each quote supported the assigned label.

**Results:** Evidence requirements modestly reduced coverage (GPT-5.2 86.2% to 84.3%, Gemini 98.3% to 92.8%, Claude 96.0% to 94.5%) by increasing abstentions and, for Gemini, invalid outputs. Conditional macro-F1 remained high but changed by model (slight gains for GPT-5.2 and Gemini, decrease for Claude). Labels were stable across repetitions (Fleiss’ kappa 0.829 to 0.969). Mechanically valid quotes occurred in 83.3% to 91.2% of runs, yet only 48.0% to 78.8% of evidence-bearing predictions were judged semantically supported. Restricting to supported predictions increased macro-F1 at the cost of lower coverage.

**Conclusion:** Substring-verifiable quotes provide an automated audit trail and enable selective, higher-trust automation when applying LLMs to biomedical text processing. However, this approach introduces new failure modes and trades coverage for verifiability in a model-dependent way.

## Introduction

Large language models (LLMs) are increasingly used for biomedical text processing and information extraction, including clinical trial matching, eligibility screening, and evidence synthesis workflows.^1–3^ In parallel, evaluations of LLMs in clinical and biomedical settings still disproportionately emphasize end-task accuracy, while transparency, verification, and trustworthiness are assessed less consistently.^4^ The purpose of this project was therefore to investigate whether a lightweight, mechanically or semantically checkable “show your work” requirement can influence the performance and improve the auditability of LLM-based eligibility classification in realistic, out-of-the-box use.

Clinical trial eligibility criteria determine which patients can enroll, shaping recruitment feasibility, representativeness, and the applicability of trial findings. In oncology, eligibility restrictions tied to disease extent are particularly consequential for both outcomes and generalizability.^5,6^ At the same time, eligibility language is frequently expressed as semantically dense free text, making automated parsing and consistent interpretation difficult.

Accordingly, a substantial literature has developed around structuring eligibility criteria into computable representations to support cohort definition and patient-trial matching. Early systems such as Criteria2Query operationalized trial criteria by converting free text into structured representations and executable database queries, with newer iterations incorporating modern LLMs to assist or semi-automate parts of this transformation.^7,8^ Complementary efforts have scaled this idea into large eligibility knowledge bases derived from ClinicalTrials.gov, enabling population-level analyses of criteria patterns and downstream informatics applications.^9^ However, across both traditional NLP and LLM-based approaches, evaluation often collapses model behavior into a single label or structured output, leaving open a central question for trust: Is each individual decision grounded in the provided text, or is it an unsupported assertion that merely happens to be correct on average?

This question is especially pressing because modern LLMs can generate fluent, confident outputs that are not reliably supported by evidence, including fabricated or erroneous bibliographic references.^10,11^ In higher-stakes clinical tasks such as medical summarization and note-generation, hallucinations and omissions have been framed as safety-relevant failure modes, motivating dedicated assessment frameworks rather than accuracy-only reporting.^12^ Recent work has also begun to operationalize “citation quality” for medical LLMs through benchmarks and automated evaluation of whether references are relevant and verifiable, underscoring both the importance and the difficulty of attribution in practice.^13^

A complementary path to trustworthy outputs is to require evidence that is directly verifiable against the input, such as extractive rationales or verbatim quotations that can be checked. In interpretability research, extractive rationales are explicitly framed as input-derived evidence that can help users judge whether a prediction is trustworthy.^14^ Similarly, in open-book question answering, models trained or prompted to provide answers alongside verified quotes from provided documents demonstrate that quote-based evidence can improve appraisal and debugging, while also illustrating that “having a quote” is not, by itself, sufficient to guarantee truth if the source is incomplete or misleading.^15^ Yet, despite growing interest in attributed and evidence-supported generation, there is limited empirical understanding of how a strict, substring-verifiable quote requirement changes LLM behavior on structured biomedical classification tasks, where the primary objective is not explanation, but reliable extraction.

In this study, we examine this gap using a structured eligibility-scope classification task on oncology randomized controlled trial (RCT) abstracts: Given abstract text alone, flagship models from three vendors must classify whether eligibility permits enrollment of patients with localized disease, metastatic disease, both, or whether eligibility scope is unclear. Each abstract is evaluated in two conditions under identical input text: A baseline run producing only a structured label, and a second run that must produce the same label plus a verbatim supporting quote drawn from the abstract, subject to a hard automatic constraint that the quote must be an exact substring after consistent whitespace normalization. This design enables annotation-free fidelity checks (format validity and quote validity), and makes it possible to measure not only correctness, but correctness under a mechanical constraint (“correct and mechanically-valid”). Furthermore, we evaluate an additional approach where a second LLM is only provided with the label and quote by the first LLM and has to assess whether the quote constitutes appropriate evidence for the assigned label.

By isolating a minimal intervention, i.e. forcing models to “show their work” via verbatim evidence, we aim to provide empirically grounded guidance on when evidence requirements improve trustworthiness, where they introduce new failure modes, and how these effects vary across vendors under realistic default settings.

## Methods

### Data selection

We reused an existing, previously annotated dataset of oncology RCT publications assembled from high-impact medical journals. The source dataset consists of 200 randomly drawn oncology RCT publications published between 2005 and 2023 across six major journals (British Medical Journal, JAMA, JAMA Oncology, The Lancet, Lancet Oncology, New England Journal of Medicine). No journal-level quotas were applied.

For the present study, the unit of analysis was the title and abstract text for each trial, reflecting realistic abstract-only screening and eligibility-parsing workflows. Full texts were not provided to the models at any stage. Trials in tumor entities where the localized versus metastatic distinction is not typically made (e.g., hematologic malignancies) were excluded, consistent with the underlying dataset construction.

### Ground-truth annotation and label mapping

Ground-truth labels were derived from the previously established manual annotation workflow for localized versus metastatic eligibility. In that workflow, trials were labeled with “LOCAL” if patients with localized or locally advanced, non-metastatic disease were eligible and with “METASTATIC” if patients with metastatic disease were eligible. Trials enrolling both populations received both labels.

The original annotation relied on title and abstract first (performed by P.W.), with full text consulted when title and abstract were inconclusive. A second author (J.W.) independently re-annotated the sampled trials using the full text, and disagreements were resolved by discussion with adjudication by a third author (D.R.Z.) if needed. In cases of discrepancy, the discussion between the annotators was based on the full-text, ensuring that each included trial had a definitive eligibility-scope label even when the abstract was incomplete. For the present study’s multi-class evaluation, we mapped the finalized label sets into four definitive eligibility-scope classes: LOCALIZED (LOCAL only), METASTATIC (METASTATIC only), BOTH (LOCAL and METASTATIC), and NEITHER.

### LLM-based classification

We evaluated three current flagship models from OpenAI, Google, and Anthropic accessed via their respective APIs using a unified local pipeline. Models were run under default vendor configurations to approximate out-of-the-box deployment. No temperature sweeps, prompt ensembling, or model-specific tuning was performed beyond what was necessary to execute the API requests. We used the model snapshots *gpt-5.2-2025-12-11*, *gemini-3-flash-preview*, and *claude-opus-4-5-20251101*.

Each trial was presented as a single-turn input consisting of the publication title followed by the abstract. Each trial was evaluated under two output conditions using identical input text. In the baseline condition, the model was instructed to output exactly one eligibility-scope label. In the evidence-required condition, the model was instructed to output the label plus a supporting verbatim quote drawn from the abstract. Baseline and evidence-required evaluations were executed as independent requests so that the evidence-required output could not condition on a prior baseline answer. The exact prompts are available from the repository.

To quantify run-to-run stability, we repeated each condition three times per trial and model under identical prompts and inputs, yielding three baseline outputs and three evidence-required outputs per trial-model combination.

The baseline system prompt required the model to output exactly one of the following labels in all caps: LOCALIZED, METASTATIC, BOTH, NEITHER or UNCLEAR. The evidence-required prompt used the same label set but additionally required a quote, returned in a fixed two-line format with a label line and a quote line. If the output from the model was UNCLEAR, no quote was required. The intent of allowing UNCLEAR was to permit an abstention when the abstract did not contain sufficient explicit information to support a stage-scope decision, even though a definitive ground truth was available from full-text-assisted human labeling.

### Mechanical output validation and verbatim evidence checking

All outputs were validated automatically. Baseline outputs were considered valid only if the returned text, after trimming whitespace, matched exactly one of the five allowed labels. Evidence-required outputs were considered format-valid only if a parsable label and a non-empty quote were present in the expected two-line format.

For evidence-required outputs, mechanical quote validity was evaluated using a strict substring-verification rule designed to be mechanically checkable. The abstract text and the model-provided quote were normalized by collapsing all whitespace runs to a single space and trimming leading and trailing whitespace. If the quote field began or ended with quotation marks, these were stripped prior to normalization. A quote was considered mechanically-valid only if the normalized quote appeared as an exact, contiguous substring of the normalized abstract. Quotes failing this check were recorded as invalid-quote regardless of label correctness.

### Semantic evidence support validation

As a second validation step, we investigated an additional approach where a second LLM was only provided with the label and quote by the first LLM and had to assess whether the quote constituted appropriate evidence for the assigned label. Each run from the initial predictions was evaluated by all three models as judge LLMs. The models were instructed to respond with either SUPPORTED or NOT SUPPORTED. The exact prompts are available from the repository.

### Endpoints and evaluation strategy

Because ground truth for all included trials was determinate, the model label UNCLEAR was treated as an abstention rather than a fourth ground-truth class. We therefore evaluated models along two complementary axes: coverage and conditional correctness.

Coverage was defined as the proportion of trials for which the model produced a non-UNCLEAR, non-invalid label. UNCLEAR frequency was summarized separately for baseline and evidence-required conditions.

Conditional performance was computed on the subset of trials for which the model produced a non-UNCLEAR label. On this answered subset, we computed accuracy and macro-averaged F1 across the four ground-truth classes (LOCALIZED, METASTATIC, BOTH, NEITHER).

For the evidence-required condition, we additionally reported the proportion of outputs that were mechanically-valid, and we defined “correct and mechanically-valid” as outputs that were both label-correct (relative to the ground truth) and mechanically-valid.

For the semantic evidence evaluation, we reported the proportion of predictions that were deemed semantically grounded by each judge LLM. In addition, we computed a macro-averaged F1 that was calculated only on semantically grounded predictions.

### Reproducibility

Run-to-run stability within each condition was quantified using Fleiss’ κ across the three repeated invocations per trial. Citation agreement across the three repeated runs was quantified using mean pairwise Jaccard similarity at the word level (number of shared words divided by the number of unique words across both quotes).

### Statistical analysis

Primary comparisons between baseline and evidence-required conditions were performed per model family. For conditional performance, comparisons were performed on the subset of trials in which the model provided non-UNCLEAR labels in the evaluated condition(s), and results were summarized using accuracy and macro-F1. Where paired hypothesis tests were used to compare accuracies between conditions, they were applied to paired trial-level outcomes using a McNemar framework after collapsing outputs into correct versus incorrect using majority voting.

All text handling, API calls, validation checks, and statistical analyses were performed in Python using a scripted pipeline analogous in structure to prior work from the same group, with all raw model outputs retained for audit and post hoc analysis.

### Ethical consideration

This study used publicly available abstracts from published clinical trials and contained no patient-level information. Therefore, ethics approval was not required.

## Results

### Inter-annotator agreement

The two annotators agreed on 191/200 trials (95.5%). Of the nine initially discrepant trials, seven were changed from “METASTATIC” (P.W.) to “BOTH” (J.W.) after full-text review, reflecting cases where only metastatic disease was mentioned in the abstract while localized/locally advanced eligibility was specified in the full text.

### Output validity and coverage

Across three repeated runs per trial, all models produced predominantly valid outputs (Table 1; Figure 1A). In the baseline condition, coverage (non-UNCLEAR, non-invalid outputs) was 86.2% for GPT-5.2, 98.3% for Gemini 3 Flash, and 96.0% for Claude Opus 4.5. Requiring verbatim evidence reduced coverage to 84.3%, 92.8%, and 94.5%, respectively. Invalid outputs remained 0.0% for GPT-5.2 in both conditions and decreased from 0.2% (baseline) to 0.0% (evidence required) for Claude Opus 4.5, but increased for Gemini 3 Flash from 1.5% (baseline) to 5.0% (evidence required). UNCLEAR rates increased under evidence requirement for all models (GPT-5.2: 13.8% to 15.7%; Gemini 3 Flash: 0.2% to 2.2%; Claude Opus 4.5: 3.8% to 5.5%).

**Figure 1.**
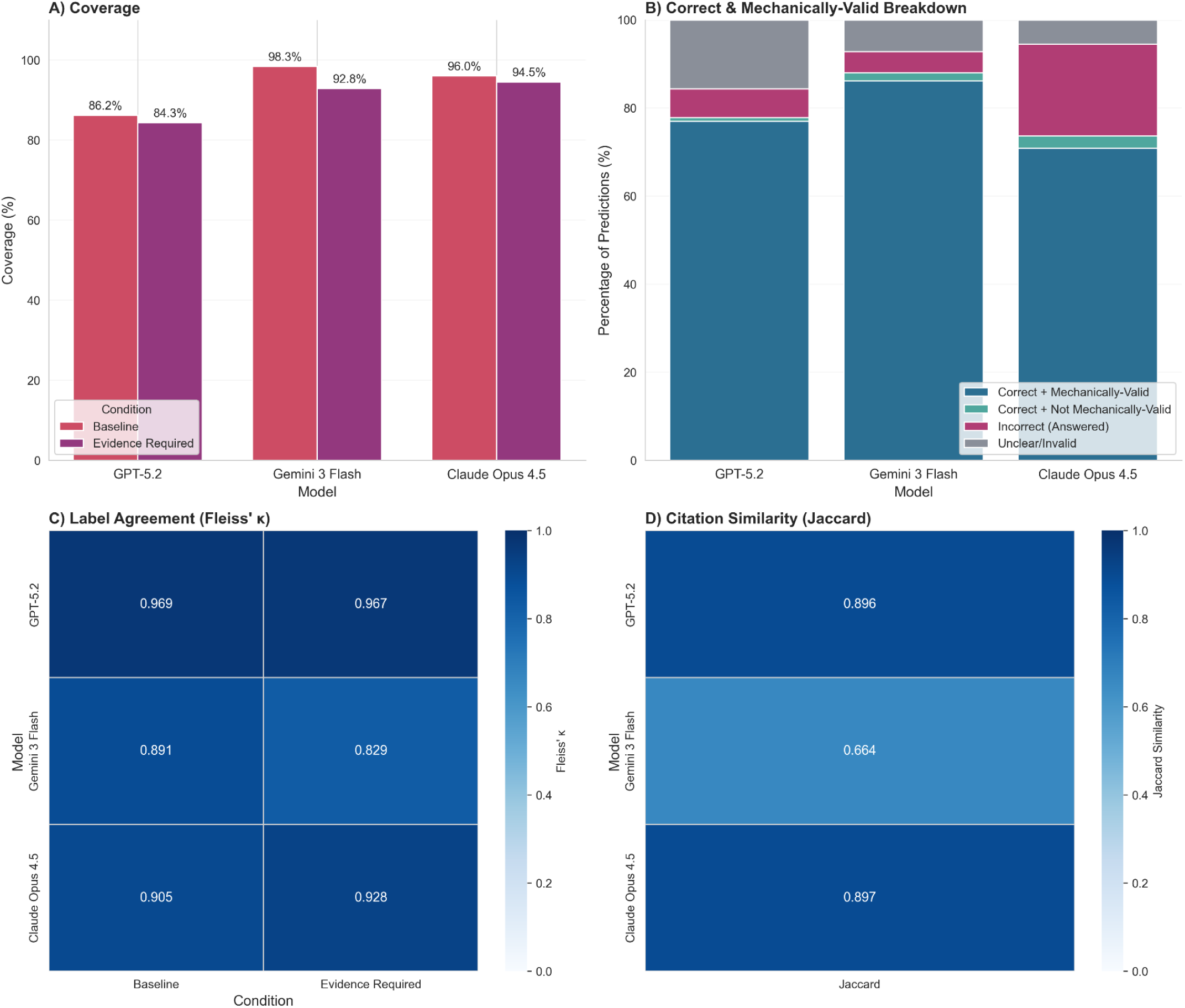
Summarized performance. A) Coverage, i.e. the percentage of non-unclear, non-invalid predictions per model and condition. B) Breakdown of predictions for the “evidence required” condition. “Correct + mechanically-valid” refers to predictions that were label-correct (relative to the ground truth) and contained a verbatim quote that was present in the abstract C) Fleiss κ for label agreement per model and condition. D) Jaccard similarity at the word level (number of shared words divided by the number of unique words across both quotes) for evidence quotes per model.

**Table 1.** Performance metrics per model and condition for GPT-5.2, Gemini 3 Flash, and Claude Opus 4.5.

| Model | Condition | Coverage [%] | Unclear [%] | Invalid [%] | Macro F1 | Fleiss' $\kappa$ | With quote [%] | With mechanically valid quote [%] | Macro F1 (mechanically-valid only) | Correct and mechanically -valid [%] | Citation Jaccard |
| --- | --- | --- | --- | --- | --- | --- | --- | --- | --- | --- | --- |
| GPT-5.2 | Baseline | 86.2 | 13.8 | 0.0 | 0.910 | 0.969 |  |  |  |  |  |
|  | Evidence Required | 84.3 | 15.7 | 0.0 | 0.916 | 0.967 | 84.3 | 83.3 | 0.917 | 77.0 | 0.896 |
| Gemini 3 Flash | Baseline | 98.3 | 0.2 | 1.5 | 0.933 | 0.891 |  |  |  |  |  |
|  | Evidence Required | 92.8 | 2.2 | 5.0 | 0.945 | 0.829 | 93.8 | 91.2 | 0.946 | 86.2 | 0.664 |
| Claude Opus 4.5 | Baseline | 96.0 | 3.8 | 0.2 | 0.828 | 0.905 |  |  |  |  |  |
|  | Evidence Required | 94.5 | 5.5 | 0.0 | 0.777 | 0.928 | 94.5 | 91.2 | 0.774 | 70.8 | 0.897 |

### Classification performance and run-to-run stability

Conditional performance (computed on non-UNCLEAR outputs) remained high for GPT-5.2 and Gemini 3 Flash under both prompting conditions (Table 1; Figure 1). Macro-averaged F1 increased slightly when evidence was required for GPT-5.2 (0.910 to 0.916) and Gemini 3 Flash (0.933 to 0.945), while it decreased for Claude Opus 4.5 (0.828 to 0.777). Label stability across the three repetitions per trial was high, with Fleiss’ κ of 0.969 versus 0.967 for GPT-5.2, 0.891 versus 0.829 for Gemini 3 Flash, and 0.905 versus 0.928 for Claude Opus 4.5 (baseline versus evidence required).

Using the trial-level majority-vote comparison, there was no statistically significant change between baseline and evidence-required accuracy for GPT-5.2 (0.785 vs 0.775; McNemar p=0.724, continuity-corrected) or Gemini 3 Flash (0.930 vs 0.910; p=0.453), whereas Claude Opus 4.5 showed a statistically significant decrease (0.785 vs 0.725; p=0.009).

### Quote stability and validation

In the evidence-required condition, a quote was returned for 84.3% of GPT-5.2 runs, 93.8% of Gemini 3 Flash runs, and 94.5% of Claude Opus 4.5 runs (Table 1). Substring-based verification confirmed mechanically valid quotes in 83.3%, 91.2%, and 91.2% of runs, respectively. When restricting evaluation to mechanically-valid quotes, macro F1 was similar to the unfiltered evidence-required results (GPT-5.2: 0.917; Gemini 3 Flash: 0.946; Claude Opus 4.5: 0.774), corresponding to “correct and mechanically-valid” rates of 77.0%, 86.2%, and 70.8% (Table 1; Figure 1B).

Quote selection was stable across repetitions for GPT-5.2 and Claude Opus 4.5 (mean word-level Jaccard similarity 0.896 and 0.897), whereas Gemini 3 Flash showed lower quote agreement (0.664), indicating greater variability in the selected evidence spans despite generally stable labels (Table 1; Figure 1D).

In the secondary “judge” evaluation, 48.0 - 78.8% of predictions with evidence were rated as semantically supported, depending on the combination of classification model and judge model (Table 2). GPT-5.2 outputs were judged supported in 73.8 - 78.8% of cases, Claude Opus 4.5 outputs in 67.2 - 73.5%, and Gemini 3 Flash outputs in 48.0 - 59.0%. On the supported subset, macro F1 ranged from 0.924 to 0.947 for GPT-5.2 predictions, 0.932 to 0.951 for Gemini 3 Flash predictions, and 0.856 to 0.917 for Claude Opus 4.5 predictions (Table 2).

**Table 2.** Performance metrics per model for a second evaluation run where models judged whether the provided evidence was appropriate to support the initial classification.

| Classification model | Judge model | Predictions deemed semantically grounded [%] | Macro F1 (semantically grounded only) |
| --- | --- | --- | --- |
| GPT-5.2 | GPT-5.2 | 73.8 | 0.924 |
|  | Gemini 3 Flash | 78.8 | 0.946 |
|  | Claude Opus 4.5 | 74.0 | 0.947 |
| Gemini 3 Flash | GPT-5.2 | 48.0 | 0.932 |
|  | Gemini 3 Flash | 59.0 | 0.951 |
|  | Claude Opus 4.5 | 49.5 | 0.946 |
| Claude Opus 4.5 | GPT-5.2 | 67.2 | 0.883 |
|  | Gemini 3 Flash | 73.5 | 0.917 |
|  | Claude Opus 4.5 | 73.2 | 0.856 |

Across models, remaining classification errors were concentrated in the BOTH class, most commonly assigned as METASTATIC, while LOCALIZED, METASTATIC, and NEITHER were generally well separated (Figure 2). Restricting analysis to mechanically-valid quotes produced near-identical confusion patterns, whereas restricting to semantically grounded outputs increased macro-F1 in the representative judge selection shown in Figure 2 (GPT-5.2: 0.92 to 0.95; Gemini 3 Flash: 0.94 to 0.95; Claude Opus 4.5: 0.78 to 0.88).

**Figure 2.**
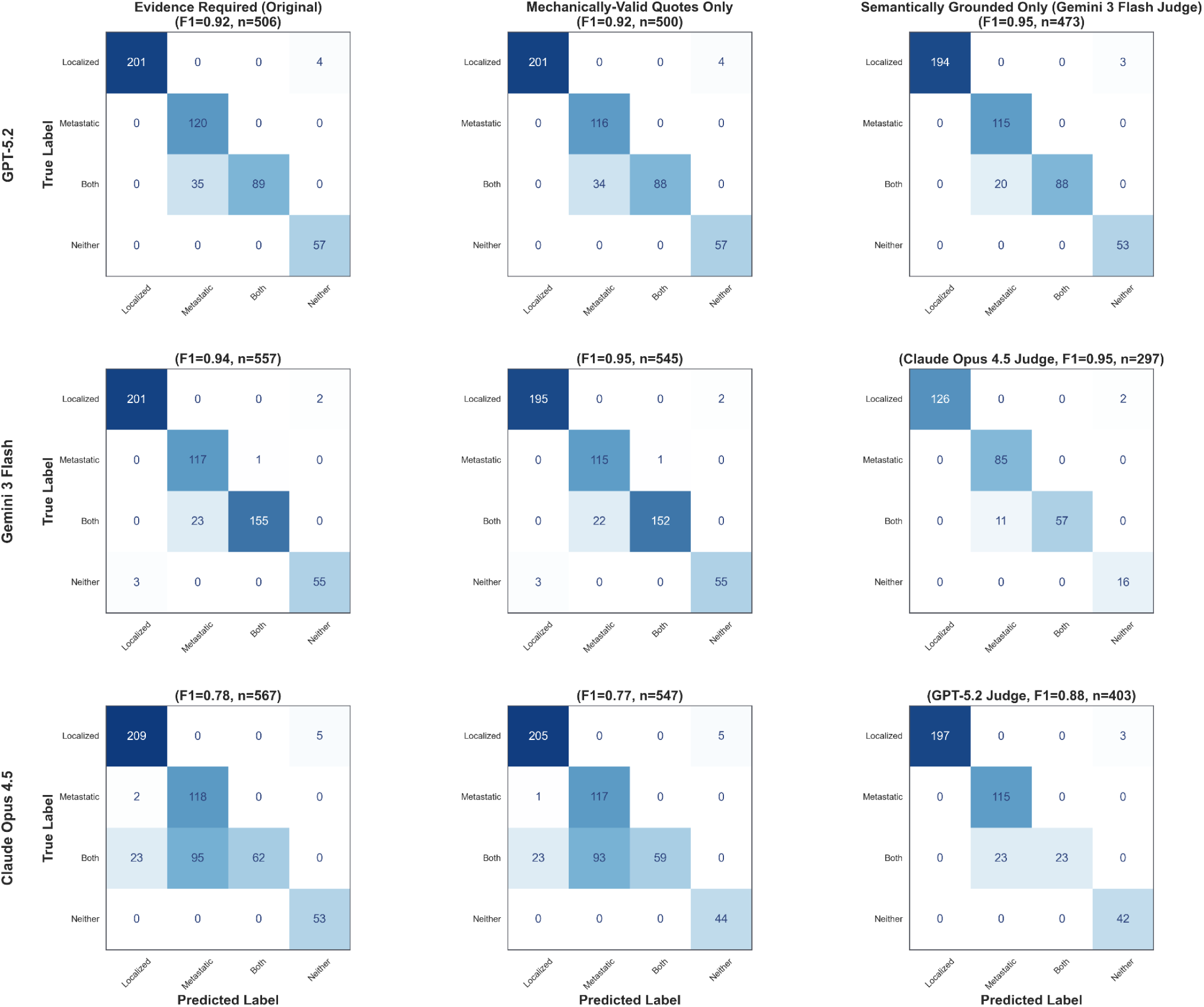
Confusion matrices per model and validation condition. All three runs per trial are plotted, except for runs that returned unclear or invalid. For the “Semantically Grounded Only” column, results of the median performing judge LLM of the respective row are depicted. From left to right, performance increases with stricter evidence requirements at the expense of reduced coverage, i.e. a lower n.

## Discussion

In this study we tested whether a lightweight “show your work” requirement - returning a label together with a verbatim supporting quote that must be an exact substring of the abstract - changes the performance and auditability of out-of-the-box LLMs for eligibility-scope classification. Across vendors, baseline conditional performance was high and run-to-run label agreement across three repeated calls was generally strong. Adding the quote constraint modestly reduced coverage by increasing abstentions and, for some outputs, introduced new failure modes. Importantly, the effect of the evidence requirement on conditional performance was not uniform: Macro-F1 increased slightly for GPT-5.2 and Gemini 3 Flash but decreased for Claude Opus 4.5, with a statistically significant decrease in majority-vote accuracy for Claude under evidence requirement. Mechanical quote validity was high but imperfect (≈ 83 - 91% across models), and evidence span stability differed substantially by vendor, with Gemini showing notably lower quote agreement across repetitions despite mostly stable labels. Finally, a secondary “judge” step suggested that mechanically valid quotes are not equivalent to semantically sufficient evidence: Depending on the classifier/judge combination, only about half to three-quarters of evidence-bearing predictions were deemed supported. However, the judge step in itself could be another method to increase the performance of LLMs on classification tasks. If only predictions with semantically grounded quotes were considered, this resulted in higher macro-F1 Scores, sometimes substantially so, however, at the expense of increasing the number of excluded predictions. For certain tasks, where a high degree of classification accuracy is required and manual review of invalid predictions is an option, automatically scoring quotes regarding semantic grounding could be a way to obtain more trustable predictions. Notably, it seems that switching models is not required as even when the initial model and the judge model were identical, the judge still declared quotes as inappropriate evidence for the assigned label.

Placed in the context of biomedical text processing more broadly, these findings speak to a persistent gap between end-task accuracy and decision-level trustworthiness. Our work operationalizes a minimal, input-grounded alternative to bibliographic citation: Rather than asking models to cite papers (which is known to be error-prone and frequently hallucinated), we require models to cite the input itself through a substring-verifiable quote. This directly addresses concerns raised by studies showing that LLMs can fabricate or corrupt bibliographic citations^10^ and that even when references exist, their relevance/supportiveness is often inconsistent and requires dedicated evaluation frameworks.^11,13^ The practical implication is that “citation” in biomedical NLP should not be treated as synonymous with “reference list generation”. Instead, enforceable, machine-checkable attribution to source text (abstracts, clinical notes, radiology reports, eligibility criteria) can be framed as a general-purpose quality control primitive for classification and extraction tasks where users need to audit why a label was produced.

At the same time, our results reinforce a core lesson from evidence-supported generation: Extractive evidence improves auditability but is not a guarantee of correctness. Open-book QA work that trains models to provide verified quotes demonstrates that evidence can support appraisal and encourages abstention when unsure, yet still leaves room for unsupported or misleading claims when the quote is incomplete or non-diagnostic.^15^ In our setting, this manifests as a clear separation between mechanical validity and semantic sufficiency: Substring-valid quotes were common, but a substantial fraction were judged not to justify the assigned label. This distinction matters for biomedical text processing pipelines that operate on heterogeneous and sometimes underspecified inputs - exactly the situation encountered not only in abstracts, but also in clinical documentation, where relevant information may be scattered, implicit, or missing. The motivation for structured, computable representations of eligibility criteria and clinical text (e.g., Criteria2Query and its LLM-enabled successors) is precisely to reduce this ambiguity, but those systems still face a trust question at the individual-decision level: Can a reviewer quickly verify that the system’s interpretation is supported by the text it saw?^7–9^ Our findings suggest that adding verbatim, checkable evidence is a feasible way to supply that audit trail without requiring new manual annotations.

The LLM-based judge step effectively transforms the pipeline into a selective prediction system, where the model can be conservative by discarding low-confidence (or weakly supported) outputs, thereby improving performance on the retained subset. This trade-off - higher accuracy (or macro-F1) at the expense of lower coverage - has direct precedent in clinical NLP: Selective prediction has been proposed and empirically evaluated as a way to improve unstructured clinical data abstraction by abstaining when uncertainty is high, with the explicit workflow assumption that abstained cases can be manually reviewed.^16^ In our approach, “uncertainty” is proxied not only by the model’s willingness to abstain (UNCLEAR), but also by whether a second model can verify that the provided quote actually supports the label. Conceptually, this resembles other reliability frameworks that trade coverage for guarantees (e.g., conformal prediction), but with an attribution-centered gating signal that is directly interpretable by humans and directly tied to the input text.^17^ For biomedical classification tasks where the cost of a wrong label is high and where human review bandwidth exists (e.g., cohort identification from notes, trial screening triage, or high-stakes phenotyping), semantic evidence scoring can therefore be positioned not merely as an evaluation add-on, but as an operational mechanism to deliver a “high-trust subset” of predictions.

The stability results add an additional, often overlooked layer to this argument. Recent methodological recommendations stress that generative AI evaluation should include repetition, because single-shot outputs can overstate reliability.^18^ In our data, labels were generally stable, but evidence spans were not equally stable across vendors, suggesting that audit trails themselves can be brittle even when headline metrics look strong. For real-world biomedical text processing, this matters because auditability is partly about reproducible justifications: If the highlighted evidence changes from run to run, downstream review interfaces and error analysis become less consistent. Concretely, this implies that evaluation of “evidence-supported classification” should routinely report not only label accuracy/F1 and quote validity, but also evidence-span stability and, ideally, risk-coverage style summaries showing how performance changes as stricter evidence filters are applied.

Several strengths support the relevance of our study. We evaluated multiple flagship models from different vendors under default settings, reflecting how LLMs are often integrated into biomedical pipelines without extensive model-specific tuning or other adaptations. The evidence requirement was deliberately designed to be mechanically verifiable, enabling fully automated detection of formatting and quote errors and allowing the study to distinguish “correct,” “correct with a quote,” and “correct with a mechanically valid quote.” In addition, the inclusion of repeated runs enabled quantification of stability and evidence reproducibility, complementing calls to incorporate stability into evaluations. Finally, the semantic judging analysis provides an initial, scalable proxy for the deeper question of whether extractive evidence is an aspect aligned with broader efforts to move beyond accuracy-only evaluation and toward safety- and reliability-aware assessment in clinical LLM applications.^4,12,19^

Key limitations remain. The abstract-only design is both a feature (ecological validity for screening workflows) and a constraint: Ground truth was established with full-text assistance, meaning that some “errors” and many abstentions may reflect appropriate behavior under incomplete evidence rather than true model failure. This is especially salient for the BOTH class, consistent with the observed pattern of residual errors and with prior observations that trial eligibility details can be restrictive, nuanced, and not fully captured in short summaries.^5,6^ The substring constraint is intentionally strict and may penalize otherwise useful evidence behavior (multi-span evidence or minor normalization differences), which could contribute to vendor-specific degradations. The judge analysis is itself limited by the known fragility of LLM-as-a-judge approaches in expert domains.

Agreement with subject-matter experts can be imperfect and variable, so semantic support rates should be interpreted as an approximate signal rather than definitive adjudication.^20^ More generally, our study focuses on one clinically meaningful label dimension in oncology RCTs and does not yet test generalization to other eligibility attributes (e.g., lab thresholds, biomarker status) or document types. Finally, all results are tied to specific model snapshots and default API behaviors. Vendor updates could change both performance and evidence compliance over time.

Several next steps would strengthen both methodological and applied impact. First, future work should systematically design and compare evidence constraints that preserve mechanical checkability while reducing brittleness, e.g., allowing multiple short spans, returning character offsets into the input text, or enforcing quoting through constrained decoding. Second, because judge reliability is itself a concern, mixed evaluation designs are warranted: Calibrate and validate judge decisions against clinician or domain-expert adjudication on a subset, and explore whether simple, task-specific heuristics (or lightweight entailment models) can complement or replace LLM judges in high-stakes settings. Third, extending beyond abstracts to full texts and to clinical documents (EHR notes, pathology reports, radiology narratives) would test whether evidence-supported classification improves real workflow outcomes such as review time, disagreement resolution, and error detection, outcomes that benchmark-style accuracy evaluations often miss.^19^ Finally, evidence-span stability should be treated as a first-class endpoint in future evaluations, because reproducible audit trails are an operational requirement in many biomedical settings.

In conclusion, requiring verbatim, mechanically verifiable evidence can make LLM-based biomedical classification outputs substantially more auditable and can encourage more conservative behavior when explicit support is lacking, but its effect on performance is vendor-dependent and mechanical validity does not guarantee semantic grounding. The judge-based semantic filter highlights a pragmatic opportunity: Evidence scoring can function as a selective prediction mechanism that produces a smaller but more accurate - and potentially more trustable - subset of outputs suitable for automation, while routing the remainder to manual review when needed.

## Data Availability

All data and code used to obtain this study's results have been uploaded to https://github.com/windisch-paul/llm_citation.

